# Motor unit firing rate modulation is more impaired during flexion synergy-driven contractions of the biceps brachii in chronic stroke

**DOI:** 10.1101/2023.11.22.23298905

**Authors:** James A. Beauchamp, Altamash S. Hassan, Laura M. McPherson, Francesco Negro, Gregory E. P. Pearcey, Mark Cummings, CJ Heckman, Julius P. A. Dewald

**Author notes:** Indicates equal contributions, co-first author (James A. Beauchamp, Altamash S. Hassan). **Contact Information of Corresponding Author:** Julius P.A. Dewald P.T., Ph.D. Department of Physical Therapy & Human Movement Sciences; Biomedical Engineering; Physical Med & Rehab. Northwestern University 645 N. Michigan Ave, Suite 1100, Room 1149 Chicago, IL 60611.

## Abstract

Following a hemiparetic stroke, individuals exhibit altered motor unit firing patterns during voluntary muscle contractions, including impairments in firing rate modulation and recruitment. These individuals also exhibit abnormal muscle coactivation through multi-joint synergies (e.g., flexion synergy). Here, we investigate whether motor unit firing activity during flexion synergy-driven contractions of the paretic biceps brachii differs from that of voluntary contractions and use these differences to predict changes in descending motor commands. To accomplish this, we characterized motor unit firing patterns of the biceps brachii in individuals with chronic hemiparetic stroke during voluntary isometric elbow flexion contractions in the paretic and non-paretic limbs, as well as during contractions driven by voluntary effort and by flexion synergy expression in the paretic limb. We observed significant reductions in motor unit firing rate modulation from the non-paretic to paretic limb (non-paretic – paretic: 0.14 pps/%MVT, 95% CI: [0.09 0.19]) that were further reduced during synergy-driven contractions (voluntary paretic – synergy driven: 0.19 pps/%MVT, 95% CI: [0.14 0.25]). Moreover, using recently developed metrics, we evaluated how a stroke-induced reliance on indirect motor pathways alters the inputs that motor units receive and revealed progressive increases in neuromodulatory and inhibitory drive to the motor pool in the paretic limb, with the changes greatest during synergy-driven contractions. These findings suggest that an interplay between heightened neuromodulatory drive and alterations in inhibitory command structure may account for the observed motor unit impairments, further illuminating underlying neural mechanisms involved in the flexion synergy and its impact on motor unit firing patterns post-stroke.

## 1. Introduction

Individuals with chronic hemiparesis following a stroke manifest multiple motor impairments, with two discordant phenomena emerging at the fundamental level of motor output (i.e., motor unit firing patterns). First, motor unit recruitment thresholds are reduced (Tang & Rymer, 1981), and the range of recruitment thresholds is compressed (Gemperline *et al*., 1995; Hu *et al*., 2015). Second, once recruited, the positive relationship between firing rate and joint torque is severely restricted (Gemperline *et al*., 1995; Mottram *et al*., 2009; Mottram *et al*., 2014). Moreover, negative “rate modulation” is often prevalent following a stroke, in which the firing rate of motor units decreases, as a function of increasing joint torque. Thus, residual motor pathways in stroke can seemingly activate motor units but are unable to modulate their discharge, forcing a greater reliance on the recruitment of higher threshold motor units to generate force.

The neuropathological mechanisms underpinning this discordant recruitment and rate modulation are likely multifactorial. At the level of the motoneuron, impaired intrinsic properties of motoneurons have been postulated post-stroke. Though this could yield the observed earlier recruitment of motor units, it has been met with conflicting evidence (McPherson *et al*., 2008b; Mottram *et al*., 2009; Mottram *et al*., 2010; McPherson *et al*., 2018b; McPherson *et al*., 2018c; Beauchamp *et al*., 2022a). Simultaneously, a shift in the primary sources of synaptic input for motor commands has been theorized. Indeed, an expanding body of evidence suggests that the disordered motor control that is observed in chronic stroke can be attributed to an increased reliance on remaining ipsilateral cortico-reticulospinal projections (Dewald *et al*., 1995; McPherson *et al*., 2018a; McPherson *et al*., 2018b; Karbasforoushan *et al*., 2019; McPherson & Dewald, 2019; Yang *et al*., 2020). Compared to the corticospinal tracts, the reticulospinal system is comprised of slower polysynaptic pathways that generate smaller excitatory post-synaptic potentials in motoneurons (Riddle *et al*., 2009; Baker, 2011), with the potential for concurrent inhibition and excitation (Koizumi *et al*., 1959). The weak potentials elicited by reticulospinal projections, and their concurrent excitatory and inhibitory commands, may ineffectively drive motoneurons and yield the impaired rate modulation observed in chronic stroke {Tian, 2021 #899}.

In addition to alterations in MU discharge characteristics, the use of cortico-reticulospinal tracts for volitional movement is thought to explain the loss of independent joint control that is observed in chronic stroke. Reticulospinal (RST) projections are relatively diffuse in nature and project to multiple motor pools, such that independent activation of distinct muscles and/or motor pools is impaired. A striking example of this is apparent in the paretic limb of individuals with chronic stroke, who often display an involuntary coupling of shoulder abduction (SABD) with elbow, wrist, and finger flexion (i.e., flexion synergy) (Twitchell, 1951; Brunnstrom, 1970; Dewald *et al*., 1995; Sukal *et al*., 2007; Lan *et al*., 2017; McPherson & Dewald, 2019). This creates a situation where excitatory drive to motoneurons of the shoulder abductors produces an involuntary activation of elbow flexor motoneurons that scales with the level of drive intended for SABD.

Given its postulated dependence on RST use in stroke, impaired rate modulation should be most prevalent during flexion synergy expression. Explicitly, during flexion synergy expression, the involuntary elbow flexion moment that is generated is likely a consequence of the diffuse RST projections to the arm motor pools, and thus primarily driven by synaptic input from the RST. If the reticulospinal system is indeed a primary mechanism of impaired motor unit rate modulation, then this aberrant discharge behavior should be more apparent in elbow flexor motor units when involuntarily activated during flexion synergy than when targeted by a voluntary effort.

In this study, we sought to investigate the relationship between changes in descending neural drive and altered motor unit firing patterns following stroke. First, we expanded on previous studies and compared voluntary contractions in the paretic and non-paretic limbs, with the capability to record from larger motor unit populations with the use of high-density surface arrays. We then quantified and compared motor unit firing rates, recruitment patterns, and rate modulation during both voluntary and synergy-driven contractions in the paretic limb. As hypothesized, we observed a reduction in rate modulation in the paretic limb and further reductions in modulation during synergy-driven contractions. Subsequently, we explore how a stroke-induced use of remaining motor pathways potentially generates these changes, using recently developed metrics informed by realistic models of motoneurons, and discuss potential underlying cellular mechanisms.

## 2. Methods

### 2.1 Participants

Eleven stroke survivors (1 female, 10 male) ranging in age from 48 to 73 (mean ± SD age: 61.91 ± 6.14) completed this study. Participants displayed a broad range of upper limb impairment levels, as quantified using the Fugl-Meyer (FM) assessment (Fugl-Meyer *et al*., 1975), with FM scores ranging from 6 to 52/66 (mean ± SD FM score: 29.90 ± 15.67). One participant did not receive a Fugl-Meyer assessment and is indicated as a dash and yellow dot in all data figure legends. For inclusion in this study stroke participants were required to have: (1) Paresis confined to one side, (2) At least one year post-stroke, (3) Absence of muscle tone abnormalities and motor or sensory impairment in the non-paretic limb, (4) Absence of severe wasting or contracture of the paretic upper limb, (5) Absence of severe cognitive or affective dysfunction, (6) Absence of severe concurrent medical problems (e.g. cardiorespiratory impairment), and (7) Absence of brainstem and cerebellar lesions as determined from clinical or radiological reports. All participants provided written and informed consent prior to participation in this study, which was approved by the Institutional Review Board of Northwestern University.

### 2.2 Experimental Apparatus

The experimental apparatus (Figure 1) is the same as used in previous work (Hassan *et al*., 2019). Participants were seated in a Biodex experimental chair (Biodex Medical Systems, Shirley, NY) and secured with shoulder and waist straps to minimize trunk movement. The participant was connected to a six degree-of-freedom load cell (JR3, Inc., Woodland, CA), using a fiberglass cast at the forearm. The arm was positioned at a shoulder abduction angle of 75°, a shoulder flexion angle of 40°, and an elbow flexion angle of 90°. Forces and torques measured at the forearm-load cell were recorded at 1024 Hz and converted into shoulder and elbow joint torques using custom MATLAB software (The MathWorks) that employed a Jacobian-based algorithm.

**Figure 1:**
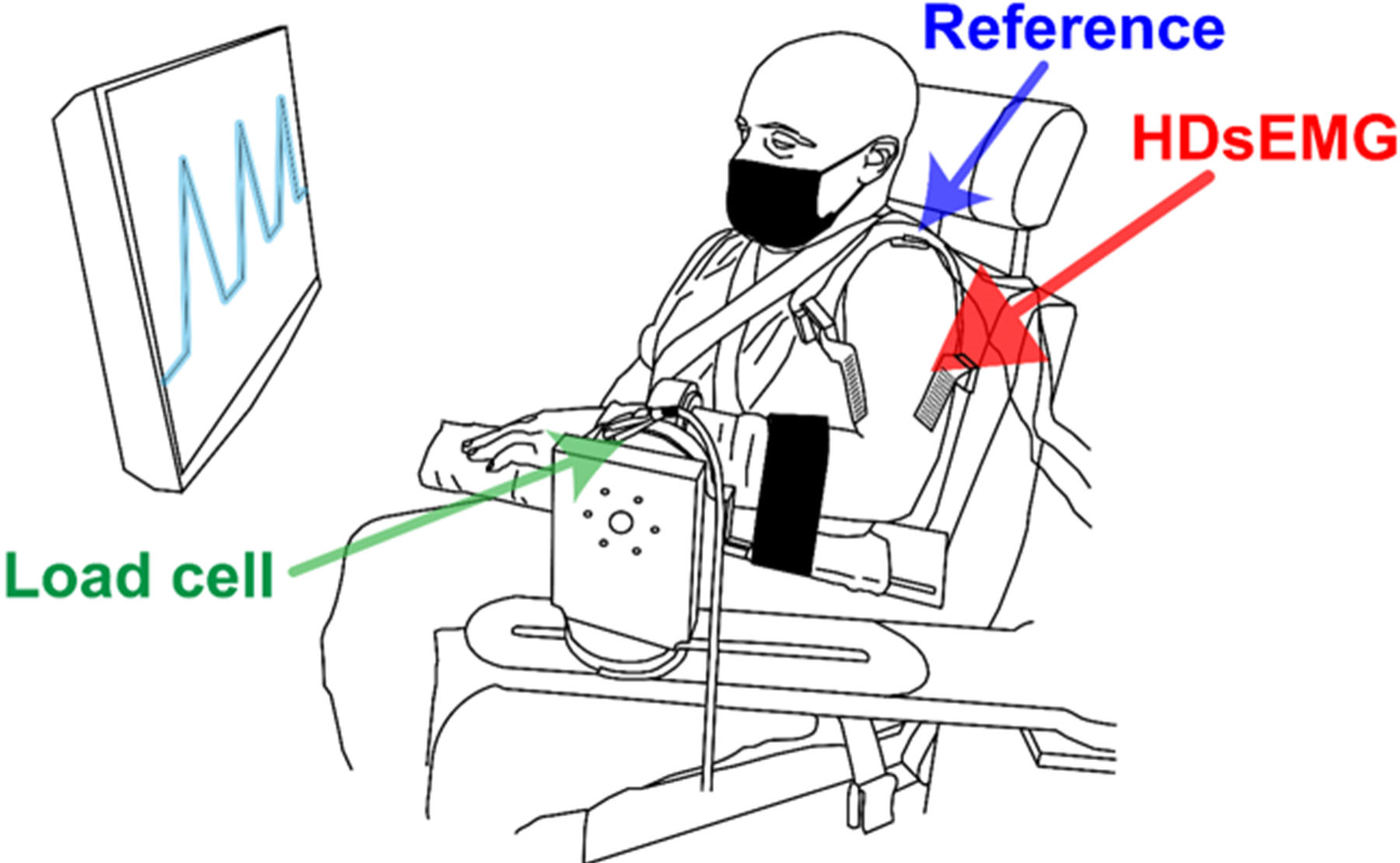
An illustration of the experimental setup. Participants are seated in a biodex chair with their forearm secured to a 6 degree-of-freedom loadcell. High-density EMG electrode grids are placed on the lateral head of the triceps brachii, and along the muscle belly of the biceps brachii. Real-time visual feedback of task performance was provided via a computer monitor in front of the participant.

Multi-channel electrode grids were placed on the surface of the biceps brachii. EMG recordings were collected in single differential mode from the 64 electrode grids with 8mm inter-electrode distance (GR08MM1305, OT Bioelettronica, Inc., Turin, IT), using a Quattrocento signal amplifier (OT Bioelettronica, Inc., Turin, IT; input range: 33mVpp). All EMG signals were amplified (x150), band-pass filtered (10-500Hz), and sampled at 2048 Hz. A 1 second TTL pulse was included as an additional channel on both the EMG and torque recordings, which allowed for off-line synchronization of the recordings.

### 2.3 Protocol

#### 2.3.1 Maximum Voluntary Torque

To normalize efforts across subjects and limbs, maximum voluntary torques (MVTs) were collected from both the paretic and non-paretic upper limbs of all participants. MVTs were collected for shoulder abduction (SABD), shoulder adduction (SADD), elbow flexion (EF), and elbow extension (EE). Participants received real-time visual feedback of their torque generation along with vocal encouragement from the experimenter. To ensure that an accurate MVT was attained, trials were repeated until the participant completed three trials in which the peak torque was within 10% of the maximum trial observed. Additionally, if the final trial showed the highest peak torque, subsequent trials were collected.

#### 2.3.2 Voluntary Submaximal Isometric EF Torque Ramps

For the experimental trials, participants generated submaximal triangular isometric EF torque ramps. Participants gradually increased their EF torque to ∼20% MVT over 10 seconds and then gradually decreased their torque back to 0% MVT over the subsequent 10 seconds. Participants received real-time visual feedback during the task, including the desired time-torque profile and a trace of their EF torque generation (Figure 1). Submaximal ramp trials consisted of three EF ramps in succession. Trials included five seconds of baseline at the beginning and end of each trial and ten seconds of rest between ramps, where no torque generation was required. Participants were given several minutes of rest between trials and several practice trials with verbal coaching to become comfortable with the task. Each participant completed five to six experimental trials that were used for subsequent analysis. Prior to motor unit analyses, all torque traces were visually inspected, and ramps with torque traces that failed to match the desired time-torque profile were removed. The submaximal EF torque ramps were collected in both the paretic and non-paretic limbs and were used for the between-limb comparisons.

#### 2.3.3 Synergy-driven Isometric Contractions

Synergy-driven contractions of the biceps brachii in the paretic limb were elicited by having participants generate SABD torque ramps. Over the course of several practice trials for each participant, we iteratively found the SABD torque level that generated ∼20% MVT EF torque via flexion synergy expression. Then, participants gradually increased SABD torque over 10 seconds to their identified target level and then gradually decreased their SABD torque over the following 10 seconds. Participants were provided with the desired time-torque profile and real-time torque feedback in the direction of SABD only. Participants completed five to eight SABD trials. Due to difficulties in obtaining synergy-driven EF contractions at exactly 20% MVT, we sought to match the exact torque level generated during the synergy-driven EF by having participants complete additional voluntary EF contractions. Participants completed five to eight additional voluntary EF trials, matching the EF torque level elicited during the SABD trials. They were provided with the desired time-torque profile and real-time torque feedback in the direction of EF for these matching trials. Each trial contained only one torque ramp, with five seconds of baseline at the beginning and end of each trial, during which no torque generation was required.

### 2.4 Data Analysis

#### 2.4.1 Motor unit decomposition

Each EMG channel was visually inspected and channels showing substantial artifacts, noise, or saturation of the A/D board were removed (typically zero to five channels were removed per trial). The remaining surface differential EMG channels were decomposed into motor unit spike trains using a convolutive blind source separation algorithm (Negro *et al*., 2016) and successive sparse deflation improvements (Martinez-Valdes *et al*., 2017). The silhouette threshold for decomposition was set to 0.85. To improve decomposition accuracy, automatic decomposition results were augmented by iteratively re-estimating the spike train and correcting for missed spikes or substantial deviations in the discharge profile (Martinez-Valdes & Negro, 2023). The instantaneous motor unit firing rates were calculated as the inverse of the inter-spike intervals of the motor unit spike trains. The instantaneous motor unit firing rates were smoothed using a 2 s Hanning window. Additionally, the rectified sum of the remaining EMG channels was used for estimates of EMG amplitude.

#### 2.4.2 Motor unit matching

To enable a more robust comparison of motor unit activity between the synergy-driven and voluntary contractions, we utilized a motor unit tracking algorithm to match individual motor units across the two contraction types. The method is based on the similarity between motor unit action potential shapes identified in different contractions. In this study, we used the normalized cross-correlation of the 2D representation of the motor unit action potentials, similar to previous studies (Martinez-Valdes *et al*., 2017; Vecchio & Farina, 2019). The method is based on the assumption that motor unit action potentials recorded by multiple electrodes over the surface of the muscle are likely to have a unique representation (Farina *et al*., 2008). The action potentials of the individual motor units were estimated by spike-triggered averaging using the decomposed firing times. In this procedure, all matches between two trials were visually inspected, and the identified motor units were classified as the same when they had a cross-correlation coefficient larger or equal to 0.80. Each matched motor unit was indexed to enable comparisons of the same unit during different contractions.

#### 2.4.3 Quantification of Rate Modulation

Motor unit firing rate modulation is here defined as the motor unit firing rate while descending excitatory input is increasing (i.e., the ascending portion of the ramp) but after the initial rapid acceleration of firing rate that occurs following motor unit recruitment. In this paper, we used EF torque as a proxy for the descending excitatory input to the biceps brachii motoneurons. We quantified the rate modulation as the slope of the linear fit of the smoothed motor unit firing rate with respect to torque, from the end of the acceleration phase to the peak of the torque ramp. Figure 2 shows the calculation of this rate modulation fit with respect to torque for a single motor unit from one stroke participant. A bilinear fit of the smoothed motor unit firing rate was used to identify the end of the acceleration phase. The transition in firing rate from during acceleration to after acceleration was determined through an iterative process that identified the best division between the first linear range and the second linear range of firing rate during the ascending limb of the torque ramp. The bilinear fit with the least error when compared to the firing rate was utilized. Motor units that were recruited after the peak in torque were excluded. Additionally, to ensure the motor units had sufficient time for post-acceleration firing, only those recruited at least three seconds before the peak torque were included in the rate modulation analyses. Since Mottram et al. (2014) reported that some motor units post-stroke demonstrate negative motor unit firing rate modulation, we calculated the incidence of negative rate modulation in our sample by determining the proportion of motor units with firing rate slope relative to torque of less than 0 pps/%MVT.

**Figure 2:**
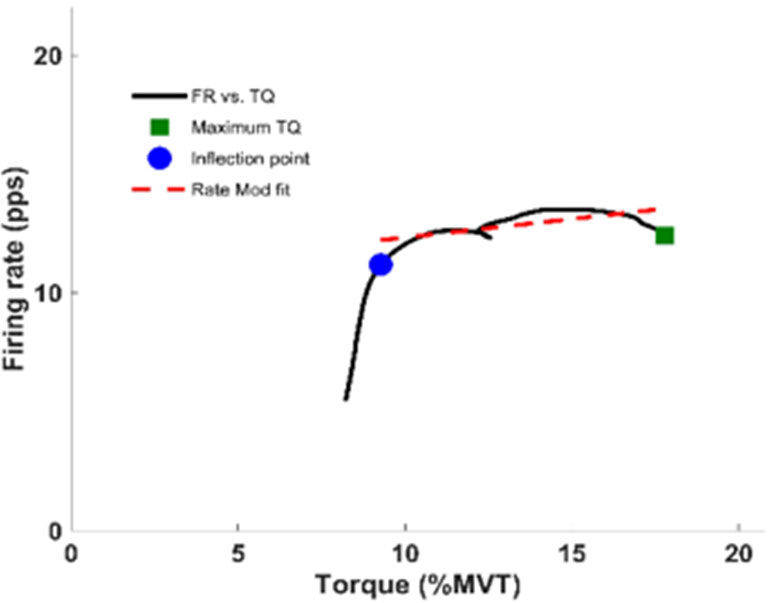
An example of the rate modulation calculation. Black line shows the motor unit firing rate plotted with respect to torque. The blue dot shows the inflection point found by the bilinear fit, identifying the end of the acceleration phase. The red dotted line is the linear fit of the post-acceleration phase, the slope of which is used as our measure of rate modulation.

Since there are multiple muscles that produce elbow flexion torque, we also wanted a measure of rate modulation that was specific to the biceps brachii alone. Therefore, we also quantified motor unit firing rate modulation with respect to the rectified sum of all EMG channels recorded from the biceps brachii. All channels without significant noise or artifacts were rectified and summed for each contraction, with this summed rectified EMG used as a proxy for descending excitatory input. However, because EMG recordings were not consistently recorded during maximum voluntary torque trials, EMG values were normalized to the largest EMG values obtained during the submaximal torque ramps for each condition (voluntary, synergy-driven, and voluntary matched to synergy-driven). Rate modulation was calculated with respect to normalized EMG (nEMG) in the same manner as it was for torque.

#### 2.4.4 Estimating the effects of an increased reliance on indirect motor pathways

To assess how a stroke-induced reliance on indirect motor pathways alters the inputs that motoneurons receive and contributes to the aberrant motor unit firing behaviors that are observed, we employed two metrics recently developed by our research group. That is, for motor units within all conditions, we quantified values of brace height and attenuation slope. Brace height represents the maximum deviation in linearity of ascending motor unit firing and attenuation slope represents the change in firing rate as a function of torque from the instance of occurrence of brace height to peak motor unit firing. Both metrics have been developed using realistic models of motoneurons and high-performance computers (i.e., supercomputers) to predict the changes in motor unit firing rate that result as a function of modulating excitatory, neuromodulatory, or inhibitory drive. We have found brace height to be a selective quantifier of neuromodulatory (i.e., monoaminergic) commands, indicating persistent inward currents in our simulations (Beauchamp *et al*., 2023; Chardon *et al*., 2023). Harmoniously, we have found attenuation slope to be a selective quantifier of the profile of inhibitory synaptic input to the motor pool (Powers & Heckman, 2017; Beauchamp *et al*., 2023). Combining these metrics grants the ability to decouple potential causative factors for the alterations in motor unit firing post stroke, a capability previously lacking in traditional approaches.

To calculate brace height and attenuation slope, we followed previously established procedures (Beauchamp *et al*., 2023; Chardon *et al*., 2023). Briefly, continuous estimates of motor unit firing were generated from decomposed discrete firing estimates with support vector regression (Beauchamp *et al*., 2022b). Braceheight was then quantified as the maximum orthogonal vector between a theoretical increase in firing and the continuous estimates of firing as a function of torque. Brace height values were normalized to the height of a right triangle whose hypotenuse represents a linear line from recruitment to peak firing. Attenuation slope was quantified as the slope of a linear line fit to motor unit firing from the instance of occurrence of brace height to peak firing. Motor units with an initial deceleration in firing rate as a function of torque or those occurring less than 1 s before peak torque were excluded from analysis.

While attenuation slope and the reported values of rate modulation slope (see Figure 2) capture similar aspects of motor unit firing behavior, we have chosen to include both. Of importance, attenuation slope is quantified from the occurrence of brace height to peak firing rate, whereas the bilinear fits used for rate modulation are quantified to peak torque. Thus, rate modulation likely yields estimates that are more behaviorally relevant and allow for characterizing the previously observed “negative rate modulation”, where the firing rate of motor units decreases as a function of increasing joint torque. In contrast attenuation slope, in its suggested form, would be unable to capture this occurrence. Instead, attenuation slope is included as a quantifier of the profile of inhibitory input, given that it has been previously validated with simulated motoneurons.

### 2.5 Statistical Analysis

We investigated the effect of limb (paretic vs. non-paretic) on the following dependent variables during voluntary contractions: peak motor unit firing rate, motor unit firing rate range, duration of motor unit firing, rate modulation slope with respect to torque, rate modulation slope with respect to nEMG, and the proportion of units exhibiting negative rate modulation. We investigated the effect of contraction type (synergy-driven vs. voluntary) on the following dependent variables during EF contractions of the paretic limb: peak torque, slope of torque ramp, RMSE of the torque trace and the slope of the torque trace, peak motor unit firing rate, motor unit firing rate range, duration of motor unit firing, rate modulation slope with respect to torque, rate modulation slope with respect to nEMG, the proportion of units exhibiting negative rate modulation, and torque at motor unit recruitment. We fit a linear mixed effects model to each dependent variable, consisting of a fixed effect of contraction type (paretic/non-paretic or voluntary/synergy) and a random effect of participant. For analysis of motor units matched across synergy-driven and voluntary contractions, motor unit index was included as a random factor. We then used estimated marginal means to compare the magnitude of difference between each condition for each metric and compute effect sizes (Cohen’s *d*).

We explored alterations in descending drive by investigating differences in brace height and attenuation slope between biceps brachii motor units during elbow flexion contractions performed in the non-paretic limb, voluntarily in the paretic limb, and involuntarily during synergy contractions. We fit a linear mixed effects model to each metric, consisting of contraction type (non-paretic voluntary, paretic voluntary, paretic synergy-induced) as a fixed effect and participant as a random effect. We then used estimated marginal means to compare the magnitude of difference between each condition for each metric.

For the comparisons of motor unit metrics, units were considered as separate data points. For the comparisons of torque traces, each trial was considered a separate data point. For the proportion of motor units exhibiting negative rate modulation, this value was determined across all trials for each study participant, and participant means were used in the linear mixed effect model.

All statistical analysis was performed with R (R Core Team, 2021). Mixed model analysis was achieved via the lme4 (Bates *et al*., 2015) package and p-values were obtained by likelihood ratio tests of the full model with the effect in question against the model without the effect in question. For main effects, this included their subsequent interaction terms. To ensure the validity of model fitting, the assumptions of linearity and normal, homoscedastic residual distributions were inspected. Effect size and estimated marginal means were employed in pairwise post-hoc testing and achieved with the emmeans package (Lenth, 2022). Cohen’s d effect size is employed and represents the difference in means as a function of the standard deviation. Estimated marginal means represent the means predicted from the statistical model for each relevant independent variable combination. They allow comparisons between the dependent variables of interest, while accounting for the appropriate fixed and/or random effects in the model. Significance was set at α = 0.05 and pairwise and multiple comparisons were corrected using Tukey’s corrections for multiple comparisons. When not reported as estimated means and confidence intervals, average values are reported as mean ± standard deviation.

## 3. Results

Decomposition from the non-paretic biceps brachii of ten stroke participants during submaximal isometric elbow flexion (EF) torque ramps yielded 435 reliable motor unit spike trains (after the assessment and cleaning procedures), with an average of 4.0 ± 2.9 motor unit spike trains per trial. In the paretic biceps brachii, 655 reliable motor unit spike trains were decomposed from eleven stroke participants, with an average yield of 5.2 ± 3.5 motor units per trial. Following the assessment and cleaning procedure, the mean silhouette values of the decomposed motor units were 0.92 ± 0.04 from the non-paretic biceps brachii, and 0.93 ± 0.04 from the paretic biceps brachii. All participants completed a minimum of nine torque ramps in both the paretic and non-paretic limb. Figure 3 shows submaximal torque ramps and decomposed motor units from both the non-paretic (A) and paretic (B) biceps brachii of one moderately impaired stroke participant.

**Figure 3:**
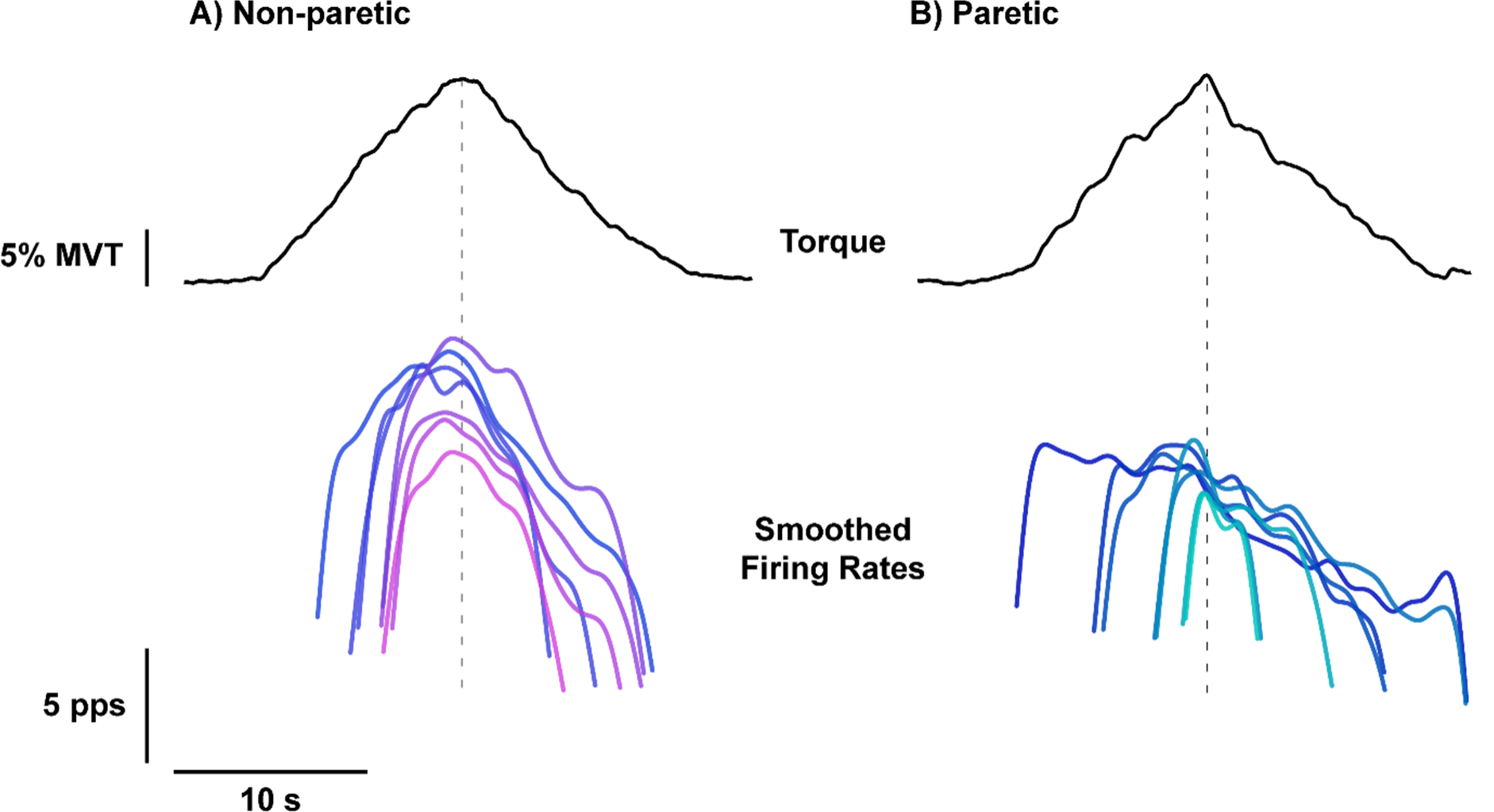
EF torque traces from the non-paretic (A) and paretic (B) limbs of one moderately impaired stroke participant are shown as black solid lines. The smoothed firing rates of decomposed motor units are shown in shades of purple for the non-paretic limb, and shades of blue for the paretic limb.

During synergy-driven contractions, decomposition yielded 394 motor unit spike trains from eleven stroke participants with an average yield of 6.4 ± 4.1 spike trains per trial. From the same eleven participants, 383 motor unit spike trains were discriminated from the biceps brachii during the matched voluntary trials, with an average yield of 5.2 ± 3.4 spike trains per trial. The silhouette values for the processed motor unit spike trains were 0.93 ± 0.03 during synergy-driven contractions and 0.93 ± 0.04 during the voluntary contractions. Each participant completed a minimum of five synergy-driven contractions and a minimum of five voluntary contractions at a torque level matched to the synergy-driven contractions.

### 3.1 Motor unit firing rate and duration in the paretic and non-paretic biceps brachii

Figure 4A shows peak firing rates of biceps brachii motor units during 20% MVT EF ramps. Colored data points and connecting lines represent participant average values from all motor units across repeated voluntary contractions. Colors indicate the Fugl-Meyer score of the participant. Notably, one participant did not receive a Fugl-Meyer assessment and is indicated as a dash and yellow dot in all figures. Black data points and vertical bars represent estimated marginal means and 95% confidence interval (95% CI), as predicted from the linear mixed effects model. Comparing the paretic and non-paretic limbs, we found limb to significantly predict peak firing rate (*χ^2^ (1) = 275.3, P < 0.0001*), with an estimated reduction of 2.7 pps (95% CI: [2.4 3.0], *d* = 1.16) in the paretic limb. Similarly, we found limb to be predictive of motor unit firing rate ranges (*χ^2^ (1) = 199.1, P < 0.0001*) with an estimated reduction of 1.7 pps (95% CI: [1.5 2.0], *d* = 0.97) in the paretic when compared to non-paretic limb (Figure 4B). Figure 4C displays the average duration of motor unit firing, which is similarly predicted by limb (*χ^2^ (1) = 117.9, P < 0.0001*) and estimated to be increased by 4.1 s (95% CI: [3.4 4.9], *d* = 0.72) in the paretic limb. This increased motor unit duration was present on both the ascending and descending limbs of the torque ramp. Limb was found to be predictive of both the duration of firing on the ascending limb (*χ^2^ (1) = 60.4, P < 0.0001*) and descending limb (*χ^2^ (1) = 100.2, P < 0.0001*) with estimated increases in the paretic limb of 1.5 s (95% CI: [1.1 1.9], *d* = 0.51) and 2.6 s (95% CI: [2.1 31], *d* = 0.66) respectively.

**Figure 4:**
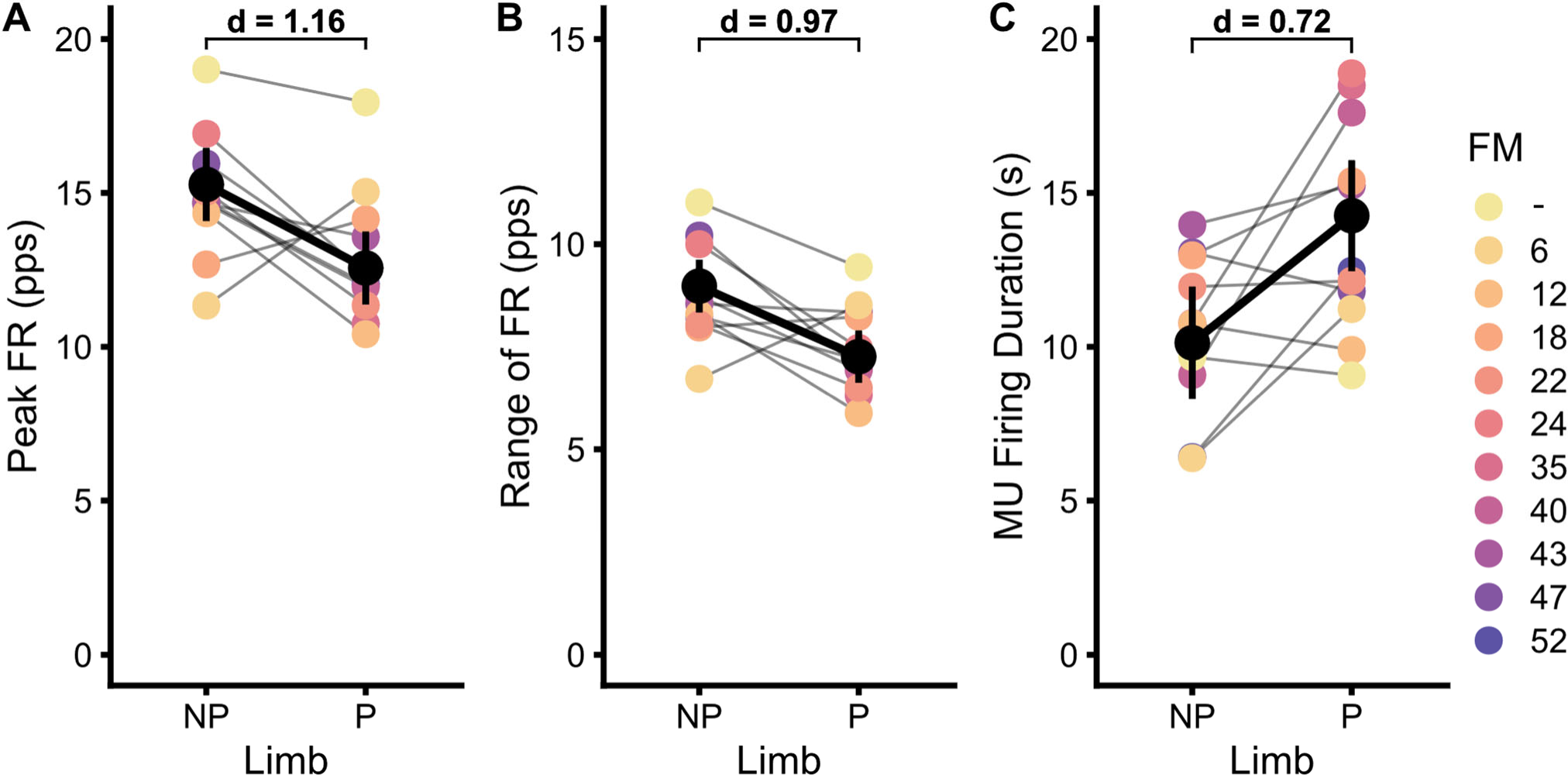
Participant averages and model estimates for peak motor unit firing rate (A), range of motor unit firing rate (B), and duration of motor unit firing rate (C) for the paretic (P) and non-paretic (NP) limb during the 20% MVT EF task. Colors indicate the Fugl-Meyer score of the participant, with connected lines representing changes in participant mean values. Black data points and vertical bars represent estimated marginal means and 95% confidence intervals (95% CI). Cohen’s d is shown for significant differences between NP and P and quantified with model estimated values.

### 3.2 Motor unit firing rate modulation in the paretic and non-paretic biceps brachii

Figure 5A shows motor unit firing rate modulation slope in the biceps brachii, with respect to torque, during ramp contractions to an effort level of 20% MVT. We found limb to predict this rate modulation (*χ^2^ (1) = 32.7, P < 0.0001*), with lower rate modulation observed in the paretic biceps by an estimated 0.14 pps/%MVT (95% CI: [0.09 0.19], *d* = 0.45). To attenuate the potential contribution of other muscles acting about the elbow joint to these observed results, we also calculated rate modulation with respect to nEMG of the biceps, shown in Figure 5B. We found limb to be predicitive of this value (*χ^2^ (1) = 8.7, P = 0.0032*), with an estimated increase of 0.028 pps/nEMG (95% CI: [0.009 0.046], *d* = 0.25). Figure 5C displays the proportion of motor units that exhibit impaired rate modulation. In accordance with Mottram et al. (2014), we found limb to be predictive of this proportion (*χ^2^ (1) = 10.3, P = 0.0013*), with greater impairments in firing rate modulation in the paretic limb than in the non-paretic limb by an estimated 0.19 (95% CI: [0.08 0.29], *d* = 1.72).

**Figure 5:**
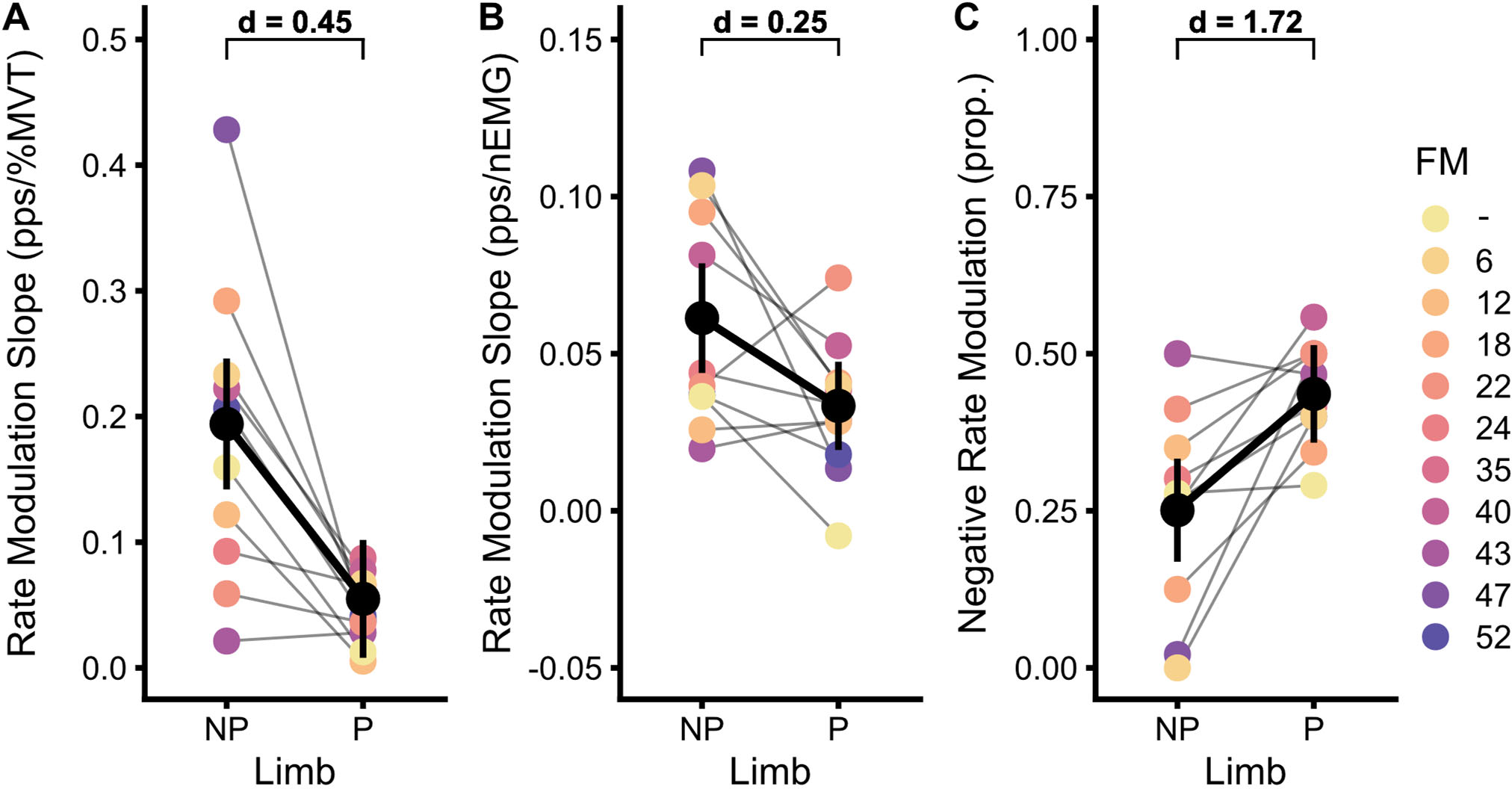
Participant averages and model estimates for motor unit firing rate modulation relative to normalized torque (A) and biceps nEMG (B) for paretic (P) and non-paretic (NP) limbs. (C) The proportion of motor units displaying negative motor unit firing rate modulation. Colors indicate the Fugl-Meyer score of the participant, with connected lines representing changes in participant mean values. Black data points and vertical bars represent estimated marginal means and 95% confidence intervals (95% CI). Cohen’s d is shown for significant differences between NP and P and quantified with model estimated values.

### 3.3 Isometric torque during synergy-driven vs voluntary contractions in the paretic limb

Stroke participants displayed a variety of EF torque behavior on the descending portion of the synergy-driven torque ramps, with some participants displaying poor relaxation and others going into elbow extension during the descending portion of the SABD task. Due to this variability, the following investigations into behavior during synergy-driven contractions are restricted to the ascending limb of the torque ramp. Participants generated peak EF torque of 20.9 ± 4.7 %MVT (range 11.6 - 28.3 %MVT) during the synergy-driven contractions. The average SABD target used to elicit the synergy-driven EF was a SABD of 30.8 ± 11.4 % MVT (range 17.9 - 45.7 %MVT).

Since the synergy-driven EF contractions were spontaneously elicited during a SABD ramp, participants did not receive real-time visual feedback of their EF torque during these tasks. To compare the motor unit activity during the synergy-driven and voluntary contractions, we first sought to ensure that the time-torque profiles during these contractions were similar. Figure 6 shows a characterization of the torque trace across all participants. Of note, one participant failed to perform the task and is shown with a dashed line in Figure 6. This participant displayed large deviations between synergy-induced contractions and the desired time-torque profile (normalized RMSE 0.85). This participant did not generate steadily increasing EF torque during synergy contractions, but instead exhibited a rapid increase in EF torque in the first few seconds that then leveled off to a constant EF torque for the rest of ascending portion of the SABD torque ramp. Due to the large discrepancies in the time-torque profile of this participant’s synergy-driven EF contractions, data from this participant were removed from all following analyses and are not included in model estimates.

**Figure 6:**
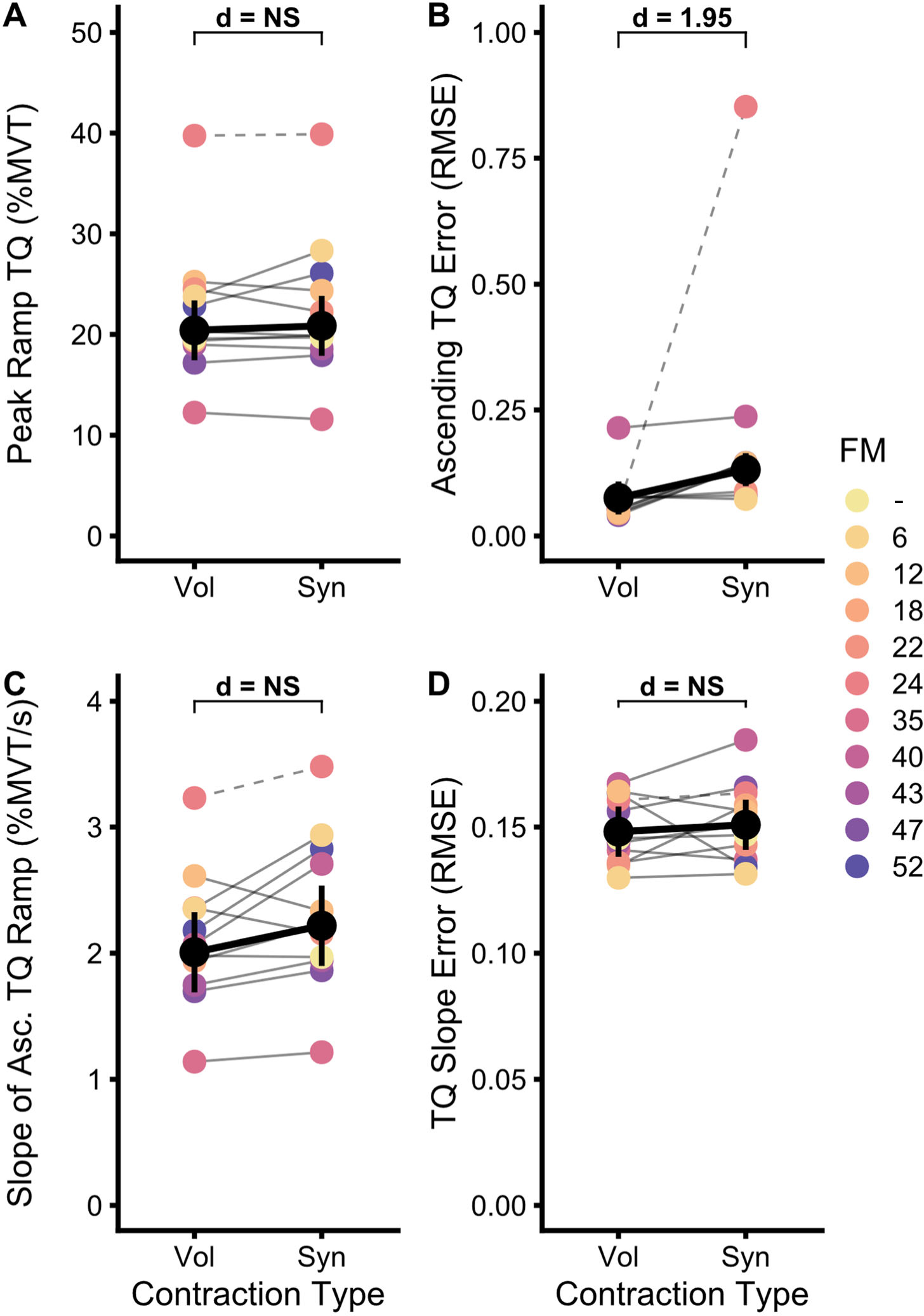
Participant averages and model estimates for peak torque (A) values during synergy-driven (Syn) and voluntary (Vol) submaximal isometric torque ramps. (B) Normalized RMSE during the ascending torque ramp, compared to the ideal time-torque profile. (C) Average torque slopes during the ascending limb of the torque ramp. (D) Normalized RMSE of the slope of the ascending limb of the torque ramp, compared to the ideal slope-time profile. Colors indicate the Fugl-Meyer score of the participant, with connected lines representing changes in participant mean values. Black data points and vertical bars represent estimated marginal means and 95% confidence intervals (95% CI). Cohen’s d is shown for significant differences between Syn and Vol and quantified with model estimated values.

The peak of the voluntary and synergy torque ramps for each participant are shown in Figure 6A. Given that the target torque levels of the voluntary contractions were matched to the peak of the synergy contractions (Voluntary: 20.4 ± 3.9 %MVT; Synergy-driven 20.9 ± 4.7 %MVT), we expectedly found no significant effect of contraction type on peak ramp torque (*χ^2^ (1) = 0.53, P = 0.4674*). Figure 6B shows the individual participant mean RMSE between the desired and performed time-torque profile during the ascending limb of the torque ramp. We found contraction type to be predictive of this error (*χ^2^ (1) = 10.6, P = 0.0011*), with an estimated increase of 0.056 RMSE (95% CI: [0.026 0.086], *d* = 1.95) in synergy-induced contractions. Although significant, practical considerations would render the magnitude of increased RMSE seen during the synergy-driven contractions small.

Looking at the slope of the torque ramps (Figure 6C), we found no effect of contraction type (*χ^2^ (1) = 3.66, P = 0.0556*). The average slope of the torque ramps is estimated as 2.22 %MVT/s (95% CI: [1.90 2.54]) in the synergy-driven contractions and 2.01 %MVT/s (95% CI: [1.69 2.33]) in the voluntary contractions. Similarly, we found contraction type to fail as a predictor for the error in torque slope (*χ^2^ (1) = 0.38, P = 0.5390*). The desired slope – time profile was a constant slope of each participant’s average peak torque during the synergy contractions divided by 10 seconds. The normalized RMSE of the torque slope is estimated as 0.151 (95% CI: [0.141 0.161]) for the synergy-contractions and 0.148 (95% CI: [0.138 0.158]) for the voluntary contractions.

### 3.4 Motor unit firing rate and duration during synergy-driven and voluntary contractions

Figure 7A displays the average peak firing rate of the paretic biceps brachii motor units during submaximal synergy-driven and voluntary EF ramp contractions at matched target torque levels. We found the contraction type (synergy or voluntary) to significantly predict max firing rates (*χ^2^ (1) = 9.10, P = 0.0026*), with these values slightly higher in synergy-driven than voluntary contractions by an estimated 0.50 pps (95% CI: [0.18 0.83], *d* = 0.24). However, we did not find contraction type to predict (*χ^2^ (1) = 1.84, P = 0.1753*) the firing rate range of motor units (Figure 7B). That said, motor units were active for a shorter duration during synergy-driven contractions, with contraction type predictive of the duration of firing (*χ^2^ (1) = 33.1, P < 0.0001*) and an estimated decrease of 2.9 s (95% CI: [1.9 3.9], *d* = 0.45) in synergy contractions. Additionally, the significant effect of contraction type on motor unit firing duration was maintained when restricting the analysis to the ascending limb of the torque ramp (*χ^2^ (1) = 48.2, P < 0.0001*). Synergy driven contractions were associated with a 1.78 s (95% CI: [1.29 2.28], *d* = 0.54) reduction in motor unit firing duration prior to peak torque, in comparison to voluntary contractions (Figure 7C).

**Figure 7:**
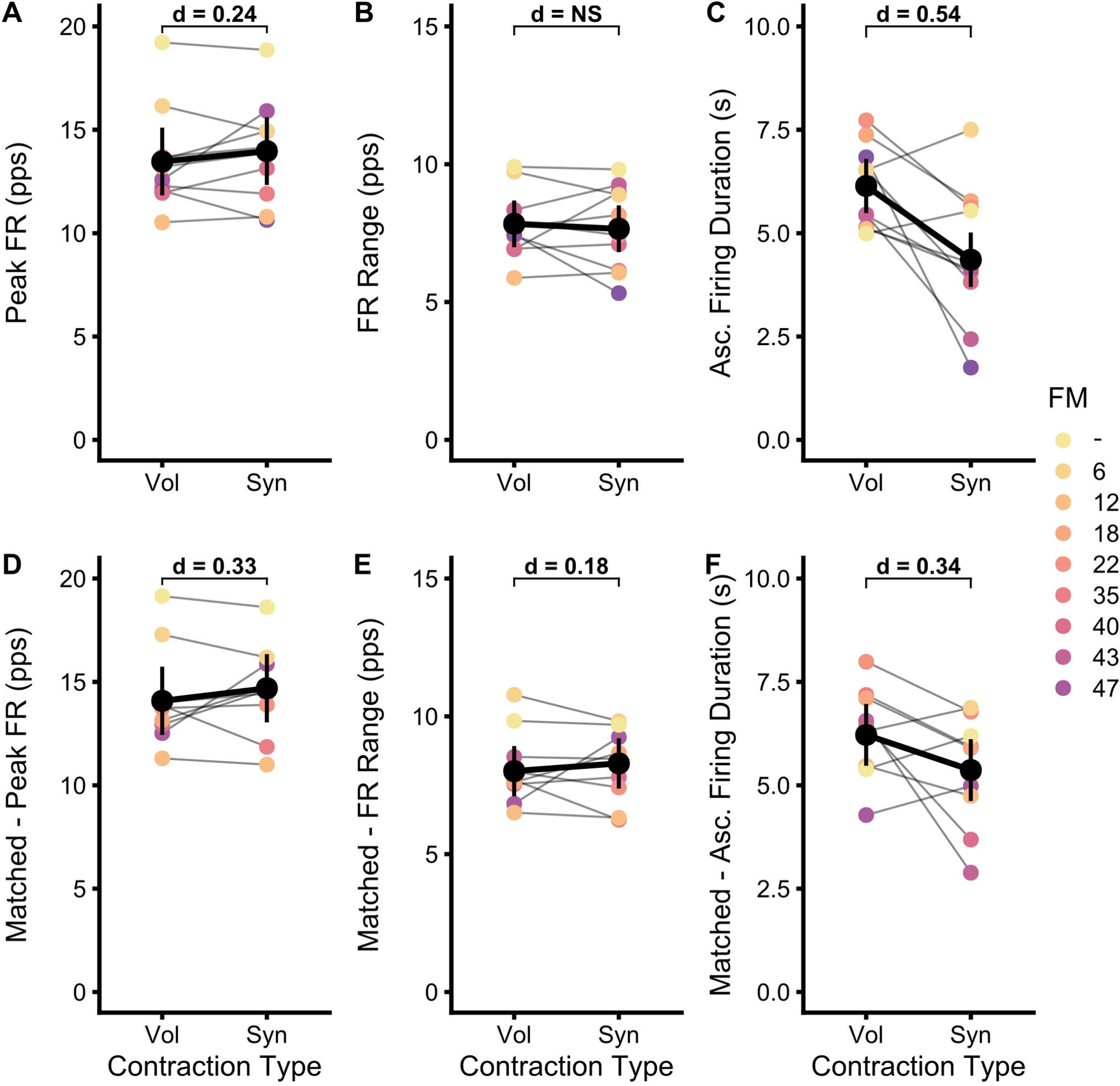
Participant averages and model estimates for peak motor unit firing rate (A), firing rate range (B), and duration of time on the ascending (asc.) phase of the torque ramps (C). These same metrics are shown in D-F for motor units matched between synergy driven (Syn) and voluntary (Vol) isometric elbow flexion contractions. Colors indicate the Fugl-Meyer score of the participant, with connected lines representing changes in participant mean values. Black data points and vertical bars represent estimated marginal means and 95% confidence intervals (95% CI). Cohen’s d is shown for significant differences between Syn and Vol and quantified with model estimated values.

When comparing matched motor units during the voluntary and synergy driven contractions, we found similar results to unmatched units. We found contraction type to be predictive of peak firing rate (*χ^2^ (1) = 26.4, P < 0.0001*), with synergy-driven contractions associated with a slight but significant increase in peak firing rate of 0.61 pps (95% CI: [0.38 0.84], *d* = 0.34). When matched, motor unit firing rate range was predicted by contraction type (*χ^2^ (1) = 7.67, P = 0.0056*) and estimated to increase during the synergy-driven contractions by 0.28 pps (95% CI: [0.08 0.48], *d* = 0.18). Furthermore, contraction type was found to be predictive of the total motor unit firing duration (*χ^2^ (1) = 22.2, P < 0.0001*) and duration on the ascending limb (*χ^2^ (1) = 27.7, P < 0.0001*) with an estimated reduction in total duration of 1.44 s (95% CI: [0.85 2.04], *d* = 0.31) and ascending duration of 0.85 s (95% CI: [0.54 1.16], *d* = 0.34) during synergy-induced contractions (Figure 7F).

### 3.5 Motor unit firing rate modulation during synergy-driven and voluntary contractions

Figure 8A shows the average motor unit firing rate modulation slope for participants during both synergy-driven and voluntary EF contractions of the paretic biceps brachii. We found the contraction type to be predictive of rate modulation slope (*χ^2^ (1) = 42.5, P < 0.0001*), with an estimated decrease of 0.19 pps/%MVT (95% CI: [0.14 0.25], *d* = 0.66) in synergy driven contraction. This reduced rate modulation slope during synergy-driven contractions was seen consistently across all 10 participants. Furthermore, all participants displayed negative firing rate modulation during the synergy-driven contractions, while only 2 of the 10 participants displayed average rate modulation that was negative during voluntary contractions. Additionally, this rate modulation as a function of nEMG in the paretic biceps during synergy-driven and voluntary contractions is shown in Figure 8B. We found contraction type to predict this rate modulation (*χ^2^ (1) = 50.2, P < 0.0001*), with an estimated decrease in synergy contractions of 0.09 pps/nEMG (95% CI: [0.07 0.12], *d* = 0.72).

**Figure 8:**
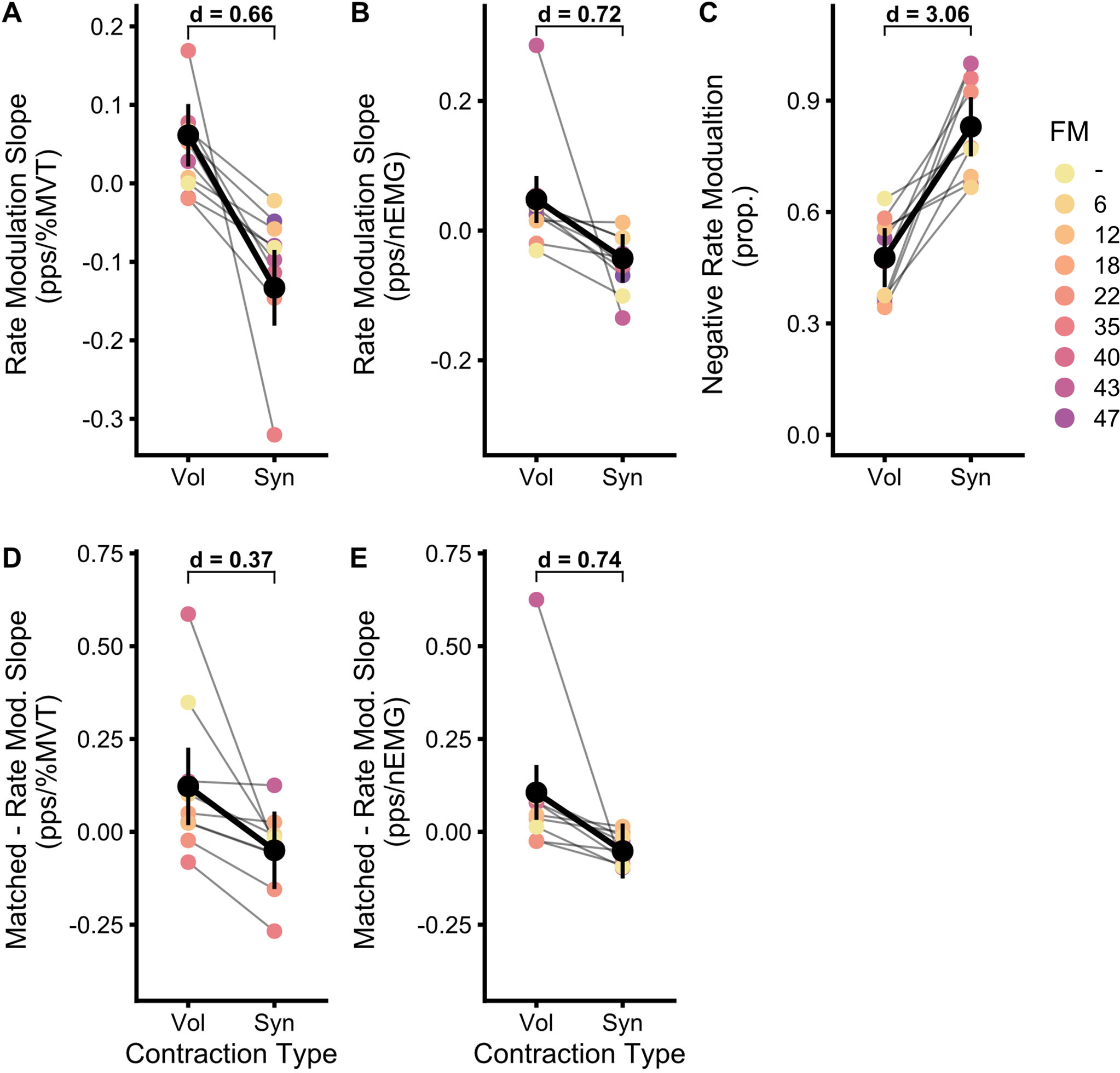
Participant averages and model estimates for motor unit firing rate modulation relative to normalized torque (A) and biceps nEMG (B) for synergy-driven (Syn) and voluntary (Vol) isometric elbow flexion torque ramps. (C) The proportion of motor units displaying negative motor unit firing rate modulation. Rate modulation metrics are shown in D-E for motor units matched between syn and vol contractions. Colors indicate the Fugl-Meyer score of the participant, with connected lines representing changes in participant mean values. Black data points and vertical bars represent estimated marginal means and 95% confidence intervals (95% CI). Cohen’s d is shown for significant differences between Syn and Vol and quantified with model estimated values.

When comparing only motor units matched between contraction types, the observed trends remain (Figure 8D-E). We found contraction type to significantly predict rate modulation with respect to both torque (*χ^2^ (1) = 22.6, P < 0.0001*) and nEMG (*χ^2^ (1) = 81.4, P < 0.0001*). For the matched units, synergy-driven contractions exhibited an estimated reduction of 0.17 pps/%MVT (95% CI: [0.10 0.24], *d* = 0.37) in rate modulation with respect to torque and a reduction of 0.16 pps/nEMG (95% CI: [0.13 0.19], *d* = 0.74) in rate modulation with respect to nEMG..

The proportion of motor units displaying impaired rate modulation during synergy-driven and voluntary contractions is shown in Figure 8C. We found contraction type to predict this proportion (*χ^2^ (1) = 24.1, P < 0.0001*), with an estimated increase of 0.35 (95% CI: [0.23 0.47], *d* = 3.06) in the proportion of motor units with negative firing rate modulation during synergy-driven contractions. This increase is strongly observed in all participants.

### 3.6 Recruitment patterns during synergy-driven and voluntary contractions

Figure 9A shows the average torque at motor unit recruitment for both synergy-driven and voluntary contractions across the entire pool of decomposed motor units (i.e., not matched between contraction types). We found contraction type to be predictive of motor unit recruitment thresholds (*χ^2^ (1) = 29.5, P < 0.0001*), and estimate average recruitment thresholds to be higher during synergy driven contractions by 2.41% MVT (95% CI: [1.55 3.27], *d* = 0.42). Notably, seven of the ten participants displayed a higher average torque at motor unit recruitment during synergy-driven contractions. Figure 9B shows the average torque at recruitment for only motor units matched between synergy-driven and voluntary contractions. Interestingly, contraction type failed to be predictive of the average torque at motor unit recruitment in the matched units (*χ^2^ (1) = 0.24, P = 0.6226*), implying that the observed differences at the population level might be due to recruitment of unique motor units.

**Figure 9:**
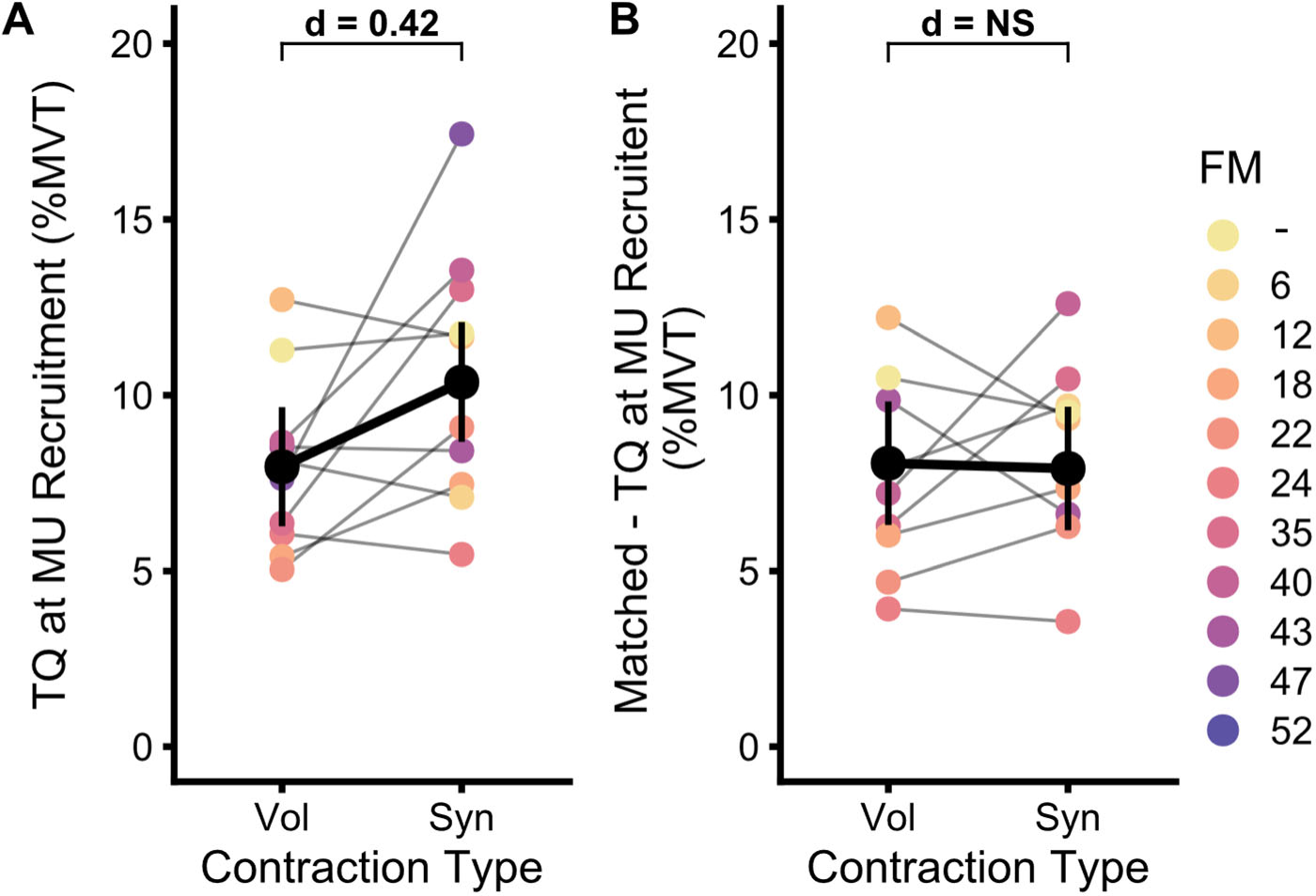
Participant averages and model estimates of average torque at motor unit recruitment for all decomposed motor units (A) or only those matched between contraction types (B). This includes synergy-driven (Syn) and voluntary isometric elbow flexion torque ramps. Colors indicate the Fugl-Meyer score of the participant, with connected lines representing changes in participant mean values. Black data points and vertical bars represent estimated marginal means and 95% confidence intervals (95% CI). Cohen’s d is shown for significant differences between Syn and Vol and quantified with model estimated values.

### 3.7 Estimating the effects of an increased reliance on indirect motor pathways during synergy-driven and voluntary contractions in the paretic upper limb

Figure 10A shows the quantification of brace height (BH) and attenuation slope (ATT) on a single motor unit (see Methods for details). Figure 10B displays the average values of brace height for isometric elbow flexion ramps generated voluntarily in the non-paretic biceps (NP), voluntarily in the paretic biceps (P-Vol), and involuntarily (synergy-driven, P-Syn) in the paretic biceps. Observing these values, we found contraction type to be predictive (*χ^2^ (2) = 23.4, P < 0.0001*), with progressive increases from voluntary non-paretic to paretic and to synergy induced. This is estimated as an increase of 4.64 %rTri (95% CI: [1.65 7.63], *d* = 0.27) from voluntary paretic to non-paretic and an increase of 7.92 %rTri (95% CI: [3.74 12.1], *d* = 0.46) from non-paretic to synergy induced.

**Figure 10:**
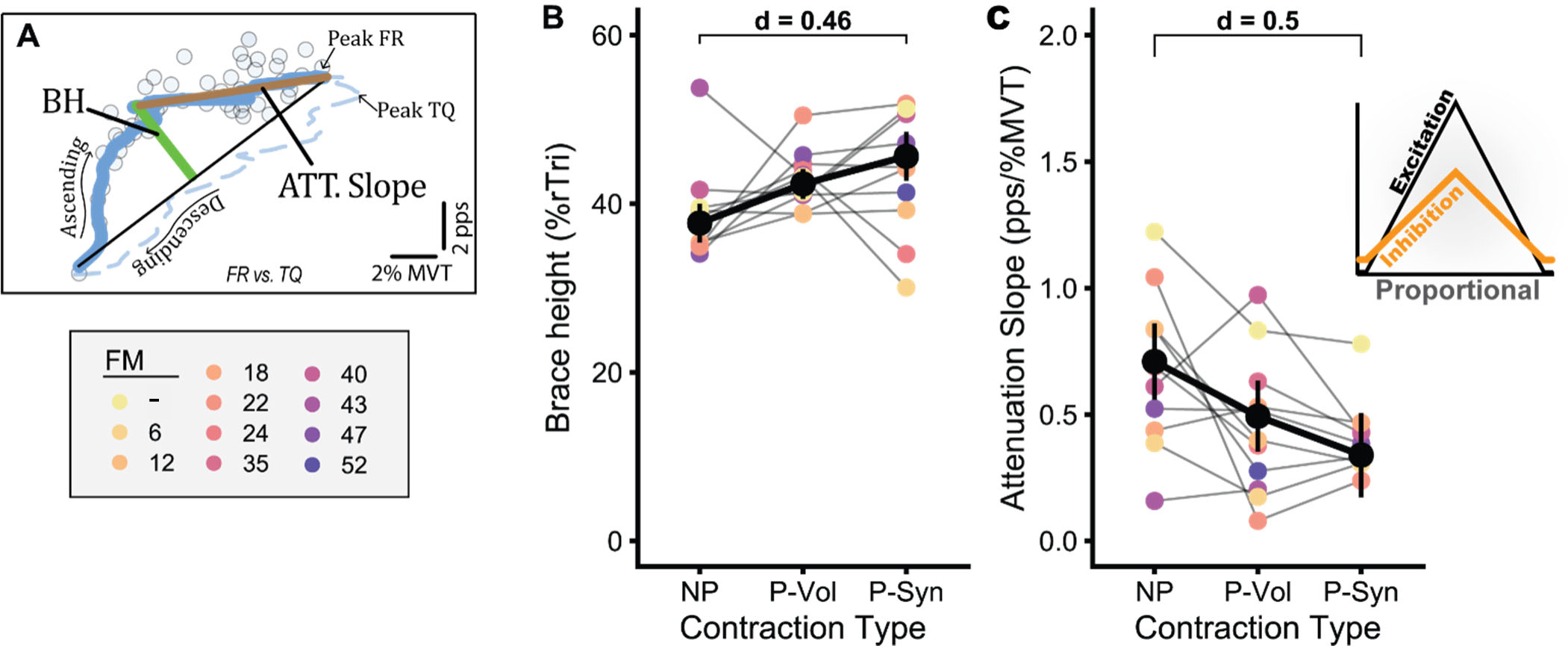
(A) Displays the quantification of brace height (BH) and attenuation slope (ATT) for a single motor unit. Participant averages and model estimates for Brace height (B) and attenuation slope (C) are shown for voluntary non-paretic (NP), voluntary paretic (P-Vol), and synergy-induced paretic (P-Syn) contractions. The inset in (C) indicates the proportional excitation-inhibition coupling pattern associated with decreases in attenuation slope. Colors indicate the Fugl-Meyer score of the participant, with connected lines representing changes in participant mean values. Black data points and vertical bars represent estimated marginal means and 95% confidence intervals (95% CI). Cohen’s d is shown for significant differences between NP and P-Syn and quantified with model estimated values.

Figure 10C displays the average value of attenuation slope for isometric elbow flexion ramps generated voluntarily in the non-paretic biceps, voluntarily in the paretic biceps, and involuntarily (synergy-driven) in the paretic biceps. We found contraction type to be predictive (*χ^2^ (2) = 25.7, P < 0.0001*) of these values with an estimated decrease of 0.216 pps/%MVT (95% CI: [0.086 0.346], *d* = 0.29) from voluntary paretic to non-paretic and 0.370 pps/%MVT (95% CI: [0.191 0.549], *d* = 0.50) from non-paretic to synergy induced. Interestingly, values of attenuation slope have been found to correspond to the excitation-inhibition coupling pattern in commands to motoneurons. The decreases observed here would be replicated by shifting the profile of inhibitory inputs to the motor pool to a pattern more proportional in nature (i.e., inhibition that scales positively with excitation). This pattern can be observed in the inset of Figure11C.

## 4. Discussion

In this study, we explored changes in biceps brachii motor unit firing patterns during voluntary isometric elbow flexion contractions in the non-paretic and paretic limb, as well as during synergy-driven and voluntarily driven isometric elbow flexion contractions. Consistent with prior research, we observed a decrease in peak motor unit firing rates and diminished firing rate modulation in the paretic limb compared to the non-paretic limb. When comparing synergy-driven and voluntary contractions, we discovered that synergy-driven contractions induced even further reductions in motor unit rate modulation, along with changes in motor unit recruitment and duration of motor unit firing. Furthermore, we examined voluntary contractions in the paretic limb, voluntary contractions in the non-paretic limb, and synergy-induced contractions in the paretic limb to assess how a stroke-induced reliance on indirect motor pathways alters the commands that motoneurons receive. Our findings indicate that both voluntary paretic and, to a greater extent, synergy-driven contractions elicit greater neuromodulatory (i.e., monoaminergic) drive and shifts in the pattern of inhibitory input received by the motor pool. Perhaps, the interplay between heightened neuromodulatory drive and a shift in inhibitory command structure, from use of indirect motor pathways, may account for the observed motor unit impairments and help explain discordant results in literature.

### 4.1 Confirmation of previously reported changes in motor unit firing behaviors post-stroke

We observed reduced motor unit peak firing rates and firing rate range in the paretic limb compared to the non-paretic limb during submaximal isometric elbow flexion contractions. (Figure 4A-B). Of note, we employed normalized target torque traces for all contractions, which yielded a lower absolute magnitude of torque in the paretic limb. This could contribute to the observed reduction in peak firing rate and firing rate range in the paretic limb. However, reduced peak motor unit firing rates have been shown previously in the paretic biceps brachii (Rosenfalck & Andreassen, 1980; Young & Mayer, 1982), even when matching absolute torque level (Gemperline *et al*., 1995). Furthermore, the observed attenuation in firing characteristics could be explained by inefficiencies in the descending motor pathways used post-stroke. Stroke survivors show a decrease in voluntary activation of the biceps brachii (Garmirian *et al*., 2019), indicating an inability to fully drive the muscle, as well as reduced integrity of the corticospinal tract (Karbasforoushan *et al*., 2019). Additionally, following a stroke, individuals might have a greater reliance on multi-synaptic cortico-reticulospinal pathways, indicated by an increase in non-linear connectivity (Yang *et al*., 2020){Tian, 2021 #899} and an increase in reticulospinal tract integrity (Karbasforoushan *et al*., 2019).

Furthermore, we observed a decrease in motor unit firing rate modulation in the paretic limb of stroke participants. Though we employed estimates from single unit firing patterns, these findings align with previous work that employed paired analysis schemes (Mottram *et al*., 2014). Across participants, we observed a consistent reduction in rate modulation slope and a large increase in the proportion of motor units that displayed impaired rate modulation (Figure 3). Previous investigations have hypothesized that reduced corticospinal input to motor units, increased persistent inward currents (PICs), and/or changes in patterns of inhibition may cause the observed reduction of rate modulation post-stroke.

Analysis of motor unit firing durations between paretic and non-paretic limbs suggests earlier recruitment of higher threshold motor units in the paretic limb and aligns with the previously observed motor unit recruitment patterns (Tang & Rymer, 1981; Gemperline *et al*., 1995),. In specific, we observed an increase in overall motor unit firing duration in the paretic limb (Figure 4C), with significant increases in firing duration on the ascending limb of the torque trace that indicate earlier motor unit recruitment. Interestingly, we also found an increased duration on the descending limb that was greater than that on the ascending limb, suggesting an increase in recruitment-derecruitment hysteresis. Motor unit hysteresis is a hallmark of monoaminergic-dependent PICs and indicates that motoneurons in the paretic limb possess larger PICs and/or receive greater neuromodulatory drive (i.e., monoaminergic) (Heckman & Enoka, 2012; Powers & Heckman, 2015).

Though greater hysteresis could indicate larger PICs and neuromodulatory drive to motoneurons post-stroke, this interpretation is clouded by conflicting findings. Though there is evidence of increased neuromodulatory drive to the paretic limb following stroke and motor impairments consistent with greater PIC-induced prolongation of motor unit firing (McPherson *et al*., 2008a; McPherson *et al*., 2018b), estimates of PICs are similar between individuals with and without stroke (Mottram *et al*., 2009). That said, previous null findings regarding estimates of PICs might be confounded by dependencies of the paired-analysis technique (i.e., ΔF) that is commonly employed. Recent and prior work from our group has highlighted the dependence of this paired analysis technique on various patterns and sources of synaptic input using realistic models of motoneurons and known sources of synaptic drive, finding ΔF to be sensitive to the pattern of inhibitory inputs to motoneurons (Powers *et al*., 2012; Beauchamp *et al*., 2023; Chardon *et al*., 2023). Given the hypothesized changes in descending drive post stroke, changes in neuromodulatory and inhibitory drive to motoneurons may produce conflicting effects on ΔF, introduce variance, and abolish the ability to detect neuromodulatory driven changes (See Figure 2a; Beauchamp et.al., 2023). Indeed, this would be consistent with our interpretation of the observed brace height and attenuation slope changes, discussed in section 4.5.

### 4.2 Comparison of elbow flexion torque traces during synergy-driven and voluntary contractions

All participants in this study generated consistent involuntary elbow flexion (EF) torque traces during shoulder abduction (SABD) ramps in the paretic limb. However, one participant (Figure 6, dashed line) exhibited diminished modulation of involuntary EF torque, displaying involuntary EF torque that was constant across SABD torque generation. As noted, this individual was removed from analysis. Furthermore, given the wide range of impairment levels, the amount of SABD necessary to elicit sufficient EF torque varied between participants. Though involuntary EF ramps across participants were consistent during the ascending portion, participants showed a variety of behavior on the descending portion of the ramps. This variation is likely due to the fact that the participants did not receive EF torque feedback and were not instructed to interfere with or modulate this spontaneously generated EF torque; their single objective was to track a linear isometric SABD ramp.

While synergy-driven EF torques were generated at a consistent level on the ascending limb of the torque ramps, the torque traces during these contractions showed a higher level of error than during the voluntary contractions (Figure 6B). This increased error during the synergy-driven contractions might be due to the lack of visual feedback provided for EF torque during these contractions. However, this error may also reflect alterations in the motor unit firing patterns (discussed below). Additionally, while this increase in torque error was significant, the torque traces during the synergy ramps showed torque increases at similar slopes as the voluntary contractions, and similar smoothness, as evidenced by comparable error in the slope of the torque ramps in both voluntary and synergy-driven contractions (Figure 6C-D). These quantitative analyses and qualitative inspection of the torque traces diminish speculation that the observed differences in motor unit firing are due to differences in EF torque-traces.

### 4.3 Differences in motor unit firing characteristics and recruitment between voluntary and synergy-driven contractions

Comparing synergy-induced and voluntary contraction, we observed a striking change in rate modulation. This can be appreciated in Figure 8 with a consistent decrease in motor unit firing rate modulation and a larger proportion of units displaying negative rate modulation in synergy-driven contractions. Of note, this proportion is estimated to be upwards of 83% during synergy induced contractions (Figure 8C), implying that over three-fourths of recruited units failed to respond to additional increases in excitatory drive upon full activation of their PIC. This highlights the severely impaired motor unit behavior present during synergy induced contractions and offers insights into causative mechanisms for post-stroke motor impairments (discussed below, section 4.5).

Furthermore, we observed a moderate shift in motor unit firing duration and recruitment patterns. Indeed, the increased torque at recruitment (Figure 9A) and shorter average firing duration on the ascending limb across the motor pool (Figure 7C), suggest that additional higher threshold motor units are recruited during synergy-driven contractions. Given that matched units did not show this shift in torque at recruitment (Figure 9B) and the difference between ascending firing duration was attenuated in matched units, the shift at the population level is likely due to recruitment of unique higher threshold units. That said, it must be noted that the alterations in torque at recruitment were inconsistent across participants, with only six of the ten participants showing an increase in average torque at recruitment. Nonetheless, this shift in recruitment pattern is likely a response to the impaired rate modulation seen in the synergy-driven contractions. As these units show severe impairments in rate coding, the only mechanism by which to increase muscle force is the recruitment of additional units.

### 4.4 Differences in motor pathways used in voluntary and synergy-driven contractions

To assess how a stroke-induced reliance on indirect motor pathways alters the inputs that motoneurons receive, we employed two metrics recently developed by our group using realistic models of spinal motoneurons and high-performance computing (i.e., supercomputers). This included the metrics coined brace heights and attenuation slope. Brace height is a quantifier of neuromodulatory (i.e., monoaminergic) commands, and attenuation slope is a quantifier of the profile of inhibitory input to the motor pool (Powers & Heckman, 2017; Beauchamp *et al*., 2023; Chardon *et al*., 2023). Combining these metrics grants the ability to decouple potential causative factors behind the alterations in motor unit firing post stroke.

We found progressive increases in values of brace height from voluntary contractions in the non-paretic and paretic limbs to synergy-driven contractions in the paretic limb. In general, these changes were moderate with the largest effect from non-paretic to synergy-driven contractions (*d* = *0*.46). Considering that the normative values of brace height in the first dorsal interosseus are ∼35 %rTri and in the tibialis anterior are ∼41 %rTri (Beauchamp *et al*., 2023), the values reported here appear reasonable and potentially elevated during synergy driven contractions (approximate values in %rTri, NP: 38, P/Vol: 42, Syn: 46). Indeed, this may represent a pathological shift in monoamines as has been theorized to occur in chronic stroke. In specific, the loss of corticofugal projections that come as a result of a stroke induced lesion to the brain is postulated to generate a relative increase in monoaminergic drive to the cord (McPherson *et al*., 2018b; McPherson *et al*., 2018c; Li *et al*., 2019). That said, comparative values of brace height in a neurologically intact biceps brachii have yet to be published as an adequate comparator.

We found progressive decreases in values of attenuation slope from voluntary contractions in the non-paretic and paretic limbs to synergy-driven contractions in the paretic limb. Of note, these changes were generally consistent across participants with a moderate effect from non-paretic to synergy-induced contractions (*d* = 0.50). Attenuation slope is a proposed quantifier of the inhibitory pattern of synaptic input received by the motor pool, whereas lower values represent an excitation-inhibition coupling that is proportional in nature (i.e., balanced, increased concurrent excitation and inhibition) and higher values represent coupling that is opposite in nature (i.e., reciprocal, increase excitation with decreased inhibition). The observed decreases in attenuation slope found in the paretic limb would indicate a shift to a more proportional pattern of inhibition (Beauchamp *et al*., 2023). Indeed, the RST has been implicated in driving excitatory and inhibitory spinal effects and recurrent inhibition has been implicated following stroke, both of which are discussed in the following section (section 4.5).

### 4.5 Speculative Mechanisms

#### 4.5.1 Reduction in corticospinal input during synergy-driven contractions

Synergy-driven elbow flexion is thought to arise from a greater reliance on the reticulospinal tracts (RST) post stroke, and their relatively diffuse projections to multiple motor pools (Dewald *et al*., 1995; Owen *et al*., 2017; Karbasforoushan *et al*., 2019; McPherson & Dewald, 2022). In short, due to the diffuse projections of the RST, excitatory drive intended for motoneurons of the shoulder abductors produces an involuntary activation of elbow flexor motoneurons. As such, descending input to the biceps during synergy-driven contractions is thought to be primarily from RST, with little corticospinal input. In contrast, during voluntary contractions of the paretic limb, individuals may use remaining corticospinal resources in concert with compensatory drive from RST pathways. Thus, progressive shifts in the balance of corticospinal and RST input from contractions in the non-paretic to voluntary paretic, and subsequently to synergy-induced contractions, may provide an explanation for the observed decrease in rate modulation. Indeed, a loss of corticospinal projections is postulated to cause an inability to fully activate motor units (Garmirian *et al*., 2019), which could yield the observed reduction in rate modulation. Additionally, increased use of cortico-RST pathways during synergy-driven contractions could further exacerbate these deficits, as the RST is primarily polysynaptic and generates smaller motoneuron EPSPs than corticospinal drive (Riddle *et al*., 2009; Baker, 2011).

#### 4.5.2 Increased neuromodulatory drive to the biceps during synergy-driven contractions

Accumulating evidence points to the role that monoaminergic drive plays in the manifestation of motor impairments in chronic hemiparetic stroke (McPherson *et al*., 2018b; McPherson *et al*., 2018c; Beauchamp *et al*., 2022a). A stroke-induced lesion of the cortex or internal capsule elicits damage to both corticospinal and cortico-reticular projections, and subsequently perturbs the balanced control of RST output. Specifically, the pontomedullary reticular formation (PMRF) receives extensive input from the cortex, exhibits many projections with contrasting influences on spinal circuitry (e.g., inhibitory dorsomedial RST, excitatory ventromedial RST), and is coupled to monoaminergic spinal cord projections (Takakusaki *et al*., 2016). Thus, loss of cortical control following a stroke may allow for greater medial RST excitability and greater monoaminergic drive to the spinal cord (Sheean, 2002; Li *et al*., 2019)(Darling *et al*., 2018; Fregosi *et al*., 2018). These monoamines (i.e., norepinephrine, serotonin) subsequently act on G-protein coupled receptors to facilitate persistent inward Na and Ca currents (PICs), which greatly amplify and prolong excitatory synaptic inputs to motoneurons.

This proposed increase in neuromodulatory drive, and thus PICs, could explain the observance of greater brace height during both voluntary and synergy-driven paretic contractions. Additionally, this may contribute to the altered recruitment characteristics and impaired rate modulation that is observed. Motoneurons with large PICs display a period of rapid acceleration, followed by a phase of rate saturation, where the unit is less sensitive to increases in descending excitatory input (Heckman & Binder, 1993; Lee & Heckman, 2000; Lee *et al*., 2003; Johnson *et al*., 2017). Thus, an increase in neuromodulatory drive could generate large PICs that saturate the motoneuron and render it less sensitive to increases in excitatory drive, yielding impaired rate modulation.

Furthermore, in part from their prolonging effect on excitatory inputs, PICs are likely critical for postural tasks (Lee & Heckman, 1998b, a), and could be higher during SABD tasks than during volitional EF tasks given the greater role of the shoulder in postural tasks. While this could explain the observed increase in brace height and motor unit characteristics during synergy contractions, given it was a SABD task, this cannot account for the observed increase in brace height from the non-paretic to paretic limb.

Of note, as recognized previously, this postulate of greater neuromodulatory drive post stroke has been met with conflicting findings. As noted, the commonly employed technique used to estimate PICs in humans (ΔF) has been found to be similar in neurologically intact individuals and individuals with stroke (Mottram *et al*., 2009). The findings of brace height and attenuation slope that we present here allow for the decoupling of neuromodulatory and inhibitory driven changes to motor unit firing, both of which have influences on ΔF (Powers *et al*., 2012; Beauchamp *et al*., 2023; Chardon *et al*., 2023). Based on the data presented here, we suggest that both a greater neuromodulatory (brace height) and more proportional inhibition (attenuation slope) contribute to the observed motor unit impairments in chronic stroke. This would exert conflicting effects on ΔF and could explain prior literature (see Figure 2A, Beauchamp et. al., 2023)

#### 4.5.3 Increased inhibitory input mediated through reticulospinal drive to Renshaw cells

Previous work has shown increased Renshaw cell excitability in individuals with spastic hemiplegia (Katz & Pierrot-Deseilligny, 1982). In specific, descending drive from the reticulospinal tract has been shown to excite both inhibitory interneurons and motoneurons (Koizumi *et al*., 1959) and have an excitatory influence on Renshaw cells (Pompeiano, 1988; Mazzocchio & Rossi, 1997). As synergy-driven contractions are likely to involve more reticulospinal drive than voluntary contractions, there may be an increase in drive to Renshaw cells during these contractions, leading to greater recurrent inhibition to motoneurons. This could explain the sizable decreases observed in rate modulation slope and explain the lower attenuation slope (i.e., proportional inhibition, concurrent excitatory and inhibitory drive).

#### 4.5.4 Possible causes for negative rate modulation in synergy-driven contractions

PIC deactivation during the synergy-driven contractions might contribute to the negative rate modulation values observed. Previous work has shown that the PIC decays within a few seconds in high threshold motor units (Lee & Heckman, 1998a). However, other than the data that we present here, there is currently no evidence that synergy drive would preferentially activate higher threshold units. Additionally, PIC de-activation might occur as a result of an increased baseline inhibition, given that PICs are quite sensitive to inhibitory inputs (Hultborn *et al*., 2003; Kuo *et al*., 2003). That said, this interpretation would require that the source of inhibition is capable of both terminating the dendritic PIC while being weak enough to spare sufficient voltage for firing at the soma.

Alternatively, an altered pattern of inhibition might generate the negative rate modulation seen in synergy-driven contractions. Indeed, this could explain the observed changes in attenuation slope and is supported by previous work showing that proportional inhibition is capable of eliciting negative rate modulation (Powers *et al*., 2012). Though causative mechanisms remain speculative, this inhibition could result from the aforementioned recurrent inhibition or an increased descending input to propriospinal neurons following stroke (Pierrot-Deseilligny & Burke, 2005), which might be due to cortico-RST projections. Propriospinal drive may utilize a combination of excitatory and inhibitory input to modulate the gain of the motoneuron pool (Chance *et al*., 2002; Berg *et al*., 2007). Further, propriospinal neurons project on to motoneurons of multiple muscles and span across joints (Alstermark *et al*., 1990), which could explain the negative rate modulation seen in the synergy-driven contractions.

### 4.6 Limitations

Several considerations and limitations must be appreciated when interpreting this work. First, these analyses were not conducted in individuals without neurological injury, instead the non-paretic limb was used as a control. However, the non-paretic limb is not entirely unaffected post-stroke, and further investigations into how these measures vary between the limbs of stroke participants and neurologically intact individuals are necessary. Additionally, the metrics of brace height and attenuation slope, though validated on motoneuron models and employed in neurologically intact participants, have not been validated on the host of alterations in descending command structure that can occur with neurologically injury. In specific, further simulations investigating changes in tonic inhibitory inputs, alterations in excitatory command profile, alterations in after-hyperpolarization period, and alterations in PIC voltage thresholds should be conducted and this data reassessed. Lastly, the possible mechanisms outlined above are necessarily based on indirect evidence and further investigation is warranted for causative explanations.

## 5. Conclusions

Here we investigated alterations in motor unit firing patterns of the biceps brachii in individuals with chronic hemiparetic stroke during voluntary isometric elbow flexion contractions in the non-paretic and paretic limbs, as well as during contractions driven by synergy and voluntary effort. Our results extend previous findings of impaired motor unit modulation in the paretic upper limb and show further impairments in rate modulation during synergy-driven contractions. Additionally, our metrics of descending commands revealed progressive increases in neuromodulatory and inhibitory drive to the motor pool from voluntary non-paretic to voluntary paretic, and subsequently to involuntary synergy induced contractions. These findings suggest that there are significant changes in neural drive during synergy-driven contractions, with greater neuromodulatory drive (i.e., monoaminergic) and concurrent inhibition (i.e., proportional) in contractions of the paretic limb that are further amplified when synergy induced. Taken together, we suggest that greater neuromodulatory drive could explain the shift in motor unit recruitment characteristics post stroke and facilitate use of the weak and diffuse reticulospinal (RST) pathways; while a more proportional pattern of inhibitory synaptic inputs (e.g., concurrent inhibition induced by Renshaw cell or RST use) could explain the observed deprecations in rate modulation.

## Data Availability

All data produced in the present study are available upon reasonable request to the authors.

## Notes

**Funding:** This work was supported by NIH grants R01HD039343 (JPAD), R01NS098509 (JPAD, CJH, AH), and T32 HD07418 (AH)

### Competing Interest Statement

The authors have declared no competing interest.

### Funding Statement

This work was supported by NIH grants R01HD039343, R01NS098509, and T32 HD07418

### Author Declarations

The Institutional Review Board of Northwestern University gave ethical approval for this work (STU00084502).

